# Analysis of Thrombotic Adverse Reactions of COVID-19 AstraZeneca Vaccine reported to EudraVigilance database

**DOI:** 10.1101/2021.03.19.21253980

**Authors:** Mansour Tobaiqy, Hajer Elkout, Katie MacLure

## Abstract

The development of safe, effective, affordable vaccines against COVID-19 remains the cornerstone to mitigating this pandemic. Early December 2020, multiple research groups had designed potential vaccines. From 11 March 2021, several European countries temporarily suspended the use of the Oxford-AstraZeneca vaccine amid reports of blood clot events and death of a vaccinated person, despite the European Medicines Agency and the World Health Organization assurance that there was no indication that vaccination was linked. This study aimed to identify and analyse the thrombotic adverse reactions associated with Oxford-AstraZeneca vaccine. This was a retrospective descriptive study using spontaneous reports submitted to the EudraVigilance database in the period from 17 February to 12 March 2021. There were 54,571 adverse reaction reports of which 28 were associated with thrombotic adverse reactions. Three fatalities were related to Pulmonary Embolism; 1 fatality to Thrombosis. With 17 million people having had the AstraZeneca vaccine, these are extremely rare events. The EMA’s Pharmacovigilance Risk Assessment Committee (18 March 2021) concluded that the vaccine was safe, effective and the benefits outweighed the risks. Conducting further analyses based on more detailed thrombotic adverse event reports, including patients’ characteristics and comorbidities, may enable assessment of the causality with higher specificity.

## 1. Introduction

The pandemic of SARS-CoV 2, the causative agent of COVID-19, poses an unprecedented challenge to world economies and community health. The development of safe, effective, affordable vaccines against COVID-19 remains the cornerstone to mitigating new viral strains in this pandemic crisis and reestablishing normality going forward. Several pharmacological therapeutics were suggested early in the pandemic for the treatment of this disease [1-3]. By early December 2020, multiple research groups had designed potential vaccines against COVID-19 with most in the early stages of approval by worldwide pharmaceutical regulatory authorities [4,5].

Chimpanzee Adenovirus encoding the SARS-CoV-2 Spike glycoprotein (ChAdOx1-S) is the Oxford-AstraZeneca vaccine designed to prevent COVID-19 infection. It was authorised by the European Medicines Agency (EMA) for use across the European Union (EU) following endorsement by the European Commission on 29 January 2021 [6]. The safety of the AstraZeneca vaccine was determined following a short-term analysis of data pooled from clinical trials that were conducted in the United Kingdom, Brazil, and South Africa [7]. Approximately 23,745 participants aged 18 years and older had been randomised and received either COVID-19 vaccine AstraZeneca or a control. The most frequently reported adverse reactions (ARs) from first vaccination were: injection site tenderness (63.7%), injection site pain (54.2%), fatigue (53.1%), headache (52.6%), malaise (44.2%), myalgia (44.0%), pyrexia (includes feverishness (33.6%) and fever >38°C (7.9%)) [6,7]. The majority of ARs were mild to moderate in severity and resolved within a short period following the vaccination [6,7]. Adverse reactions to second vaccinations were of the same nature but ‘milder and less frequent’ [6].

From 11 March 2021, several European countries (including Denmark, France, Italy, Latvia, Norway, Spain, Sweden and The Netherlands) temporarily suspended the use of the Oxford-AstraZeneca COVID-19 vaccine amid reports of blood clot events and death of a vaccinated person [8]. Within a week, some 18 countries worldwide had suspended use of the vaccine. This precautionary measure was taken despite the European Medicines Agency (EMA) and the World Health Organization assurance that there was no indication that vaccination was linked to thromboembolic events [4,6]. According to the AstraZeneca COVID-19 vaccine summary of product characteristics (for healthcare professionals), blood and lymphatic system disorders as adverse reactions were uncommon, however, no details on the occurrences and frequencies of these reactions were available at this time [9]. AstraZeneca emphasised the safety of their vaccine stating the blood clot prevalence was ‘much lower than would be expected to occur naturally in a general population of this size [9,10].

Traditionally, monitoring of vaccine safety after licensure is subject to a combination of passive and active surveillance. The passive surveillance system is the basis of pharmacovigilance, which comprises the databases into which spontaneous reports of suspected adverse drug reactions (ADRs) and adverse events following immunisation (AEFI) from physicians and patients are collected [11]. EudraVigilance (EV) is a passive pharmacovigilance system for collecting, managing and analysing suspected ADRs and AEFI reports for medicines approved in the EU and it is operated by EMA [11-13].

Monitoring of safety reports of drugs and vaccines is critical because clinical trials are designed to assess primarily the efficacy; safety is typically a secondary objective. These trials are able to identify only common adverse events such as local and systemic reactions related to the immunogenicity of the vaccine that occurs shortly after administration. The trials are unlikely to detect rare adverse events that occur with trials in larger populations or delayed reactions that occur a long time after use [7]. Furthermore, currently approved and still being investigated COVID-19 vaccines are using novel and unlicensed technologies [12-14].

Therefore, this study aimed to identify and analyse the thrombotic adverse reactions associated with Oxford-AstraZeneca vaccine following its temporary suspension in several EU countries using the European EV database.

## 2. Materials & Methods

This was a retrospective descriptive study using spontaneous reports submitted to the EV database in the period from 17 February to 12 March 2021. Data were extracted from the line listing section of reports submitted to the EV in relation to Covid-19 Vaccine AstraZeneca (ChAdOx1-S) [14]. The following search terms were applied regardless of age group, gender, geographical area, reporter or level of seriousness: Thrombosis; Embolism; Thromboembolism; Embolic and Thrombotic [15]. Data were cleaned, tabulated and presented according to the type of thrombosis and clinical outcome, by gender and age.

Ethical consideration: Open access data were used therefore no access authorisation was requested. The access policy of EMA states that *“ No authorisation for accessing the ICSR (Level 1) data set by means of the adrreports*.*eu portal is required i*.*e. all academic researchers can access adverse reaction data of interest. “* [14].

## 3. Results

Overall, there were 54,571 adverse reaction reports for ChAdOx1-S reported to the EV database. The total number of thromboembolic reports was 28 of which 19 (67%) were submitted by healthcare professionals. More than half (n=16; 57%) of the reports pertained to people aged over 85 years; 13 (47%) reports were from within the EU (Tables 1 and 2).

**Table 1:** Summary of demographics of EV database reported Astra-Zeneca vaccine ADR events (N=28)

| Variable | Level | n (%) |
| --- | --- | --- |
| Gender | Male | 9 (32.1) |
|  | Female | 19 (67.9) |
| Reporter profession | Healthcare Professional | 19 (67.9) |
|  | Non Healthcare Professional | 9 (32.1) |
| Geographical area | EU | 13 (46.4) |
|  | Non EU | 15 (53.6) |
| Age group of patients | 18-64 Years | 17 (60.7) |
|  | 65-85 Years | 8 (28.6) |
|  | More than 85 Years | 3 (10.7) |

**Table 2.** Detail of EV database reported AstraZeneca vaccine ADRs (N=28)

| Receipt Date | Primary Source Qualification | Primary Source Country | Age Group | Sex |
| --- | --- | --- | --- | --- |
| 17/02/2021 | Healthcare Professional | Non EU | 18-64 Years | Male |
| 18/02/2021 | Healthcare Professional | Non EU | More than 85 Years | Female |
| 23/02/2021 | Healthcare Professional | Non EU | 65-85 Years | Male |
| 02/03/2021 | Healthcare Professional | Non EU | 65-85 Years | Female |
| 03/03/2021 | Healthcare Professional | Non EU | 18-64 Years | Female |
| 06/03/2021 | Non Healthcare Professional | Non EU | 65-85 Years | Female |
| 06/03/2021 | Healthcare Professional | Non EU | More than 85 Years | Female |
| 06/03/2021 | Non Healthcare Professional | Non EU | 65-85 Years | Female |
| 06/03/2021 | Healthcare Professional | Non EU | More than 85 Years | Female |
| 06/03/2021 | Non Healthcare Professional | Non EU | 65-85 Years | Male |
| 06/03/2021 | Healthcare Professional | Non EU | 65-85 Years | Male |
| 07/03/2021 | Non Healthcare Professional | Non EU | 18-64 Years | Female |
| 07/03/2021 | Healthcare Professional | Non EU | 18-64 Years | Female |
| 07/03/2021 | Healthcare Professional | Non EU | 65-85 Years | Female |
| 07/03/2021 | Healthcare Professional | Non EU | 65-85 Years | Male |
| 08/03/2021 | Healthcare Professional | EU | 18-64 Years | Female |
| 08/03/2021 | Healthcare Professional | EU | 65-85 Years | Female |
| 09/03/2021 | Healthcare Professional | EU | 18-64 Years | Female |
| 09/03/2021 | Healthcare Professional | EU | 18-64 Years | Male |
| 10/03/2021 | Healthcare Professional | EU | 18-64 Years | Female |
| 10/03/2021 | Non Healthcare Professional | EU | 18-64 Years | Male |
| 10/03/2021 | Non Healthcare Professional | EU | 18-64 Years | Female |
| 10/03/2021 | Healthcare Professional | EU | 18-64 Years | Female |
| 11/03/2021 | Healthcare Professional | EU | 18-64 Years | Male |
| 11/03/2021 | Non Healthcare Professional | EU | 18-64 Years | Female |
| 11/03/2021 | Non Healthcare Professional | EU | 18-64 Years | Female |
| 12/03/2021 | Healthcare Professional | EU | 18-64 Years | Female |
| 12/03/2021 | Non Healthcare Professional | EU | 18-64 Years | Male |

Of the six cases that reported pulmonary embolism, 2 had a fatal outcome, both in females, one in each of the 18-64 years and over 85 years of age groups. Amongst men, there was one fatality following thrombosis in a patient aged 18-64 years of age. Table 3 (female; n=19) and Table 4 (male; n=9) illustrate the type of reported thromboembolic reaction along with the clinical outcome of cases by age group (Tables 3 and 4).

**Table 3:**
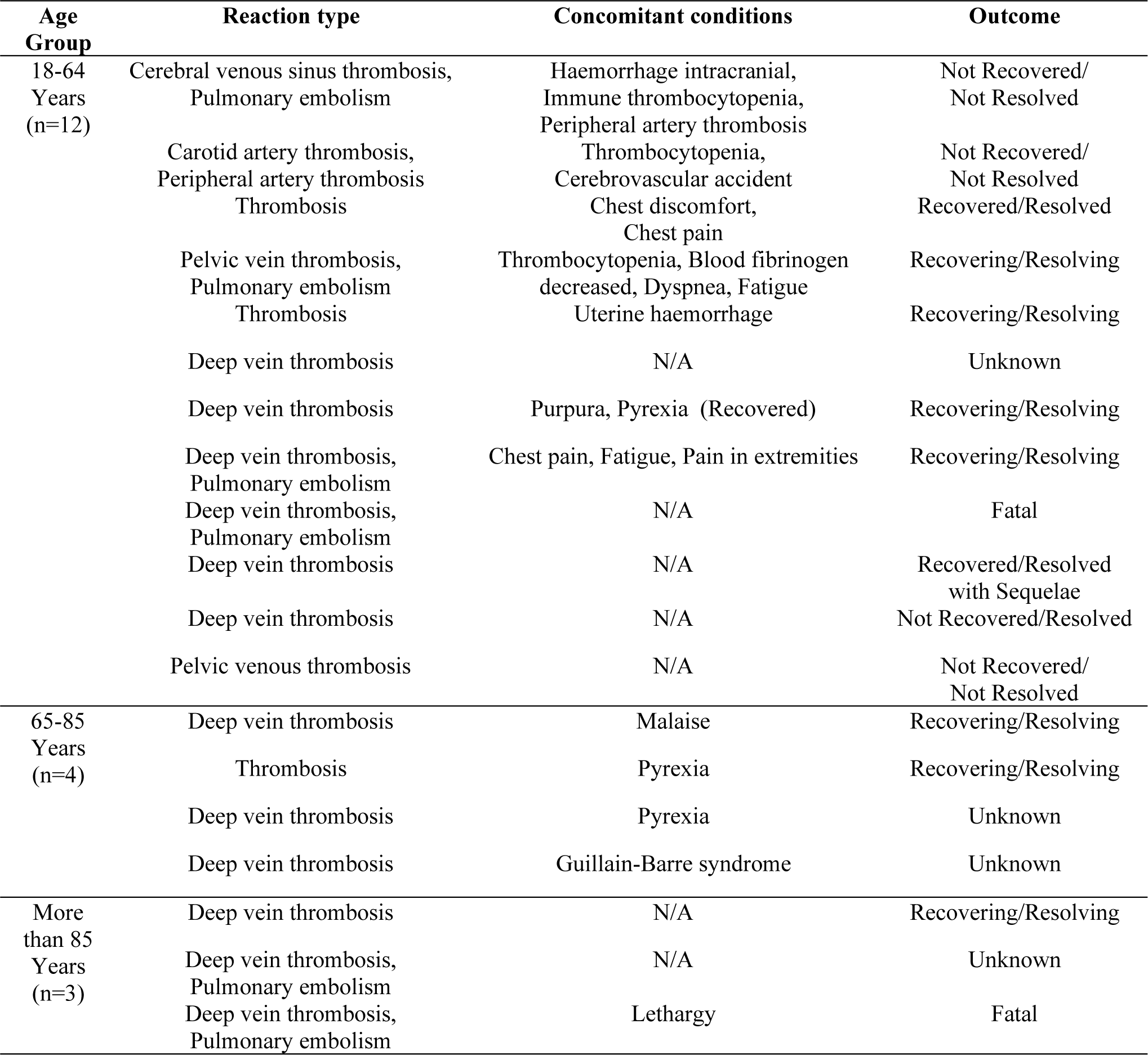
Type of thromboembolic reaction and clinical outcome for female cases by age group (N=19/28)

**Table 4:** Type of thromboembolic reaction and clinical outcome for male cases by age group (N=9/28)

| Age Group | Reaction | Concomitant conditions | Outcome |
| --- | --- | --- | --- |
| 18-64<br>Years | Thrombophlebitis | Pain | Recovering/Resolving |
|  | Thrombosis | Flu-like illness | Recovering/Resolving |
|  | Thrombosis | Arthralgia, Thrombocytopenia,<br>Pain in extremities, pyrexia | Fatal |
|  | Deep vein thrombosis | N/A | Not Recovered/Not Resolved |
|  | Deep vein thrombosis | N/A | Recovering/Resolving |
| 65-85<br>Years | Thrombophlebitis | N/A | Not Recovered/Not Resolved |
|  | Deep vein thrombosis | N/A | Recovering/Resolving |
|  | Thrombosis | N/A | Not Recovered/Not Resolved |
|  | Deep vein thrombosis | Cellulitis, Lymphadenitis | Unknown |

## 4. Discussion

To date, 17 million people in the EU and UK have received at least their first dose of the AstraZeneca vaccine [X]. This study identified 28 thrombotic adverse events linked to the use of AstraZeneca vaccine via the EV database, three of which were fatal outcomes of PE.

Pulmonary embolism (PE) and deep vein thrombosis (DVT) have usually been associated with multiple causative factors. These can be either hereditary or naturally occurring causes including cancer, advanced age, trauma, smoking, inherited or acquired thrombophilic states, previous thromboembolism, and hospitalisation for congestive heart failure or acute exacerbation chronic obstructive pulmonary disease (COPD). PE and DVT are typically associated with morbidity and mortality. Their natural incidence ranges from 56 events per 100,000 persons to 182 events per 100,000 persons which should be considered against the three deaths from PE in 17 million following vaccination [16,17]. A recent single study from Croatia reported an increase in prevalence of ‘combined DVT with PE but not with isolated PE or isolated DVT’ [18]. The authors noted from their comparison of non-Covid-19 patients in matched 8-month periods in 2019 pre- and in 2020 during the pandemic a significantly older age group admitted to hospital (60.8 ± 17.2 years v. 68.5± 16.8 years) [18]. It was postulated that pandemic lockdowns restriction on physical activity may be the cause.

In Europe, a rare disease (AE in this scenario) is defined when it affects five in 10,000 people (prevalence: 500/million). Ultra or very rare AE is defined as when it affects one in 50,000 people or 20/million. With a simple calculation of 28 thrombotic events reported in relation to the AstraZeneca vaccine for 17 million people who had the vaccine these would be cautiously labelled as extremely rare events [19].

Notably, It would be challenging and difficult to determine the causality and link the thrombotic events to AstraZeneca vaccine using spontaneous reports. Also, to apply statistical calculations, could be misleading because of the very limited safety and clinical data on the use of vaccine in the people affected, together with under-reporting and the low numbers of exposed individuals. This situation is further complicated by different confounding factors such as the indications and the wide spectrum of unknown comorbidities of patients [20].

This study also identified approximately double the occurrence of potential thrombotic events reported in females (n=19) than males (n=9). Thrombotic events in younger females are typically associated with ovarian hyperstimulation syndrome (OHSS) following reproductive treatment. Although the exact mechanism of developing thrombosis is unknown, it has been postulated that coagulation factors can result in high levels of estradiol hormone. Other specific factors for developing VTE during pregnancy including caesarean delivery, history of prior VTE, family history of VTE, inherited or acquired thrombophilia, obesity, older maternal age and prolonged immobilization [21,24].

Additionally, and according to a systematic review and network meta-analysis study, all combined oral contraceptives were associated with an increased risk of venous thrombosis, depending on both the progestogen used and the dose of ethinylestradiol [25].

Currently, the UK Joint Committee on Vaccination and Immunisation (JCVI) advised that vaccination in pregnancy should be considered in women who are frontline health or social care workers or have underlying conditions that put them at very high risk of being infected with, transmitting or experiencing serious complications of COVID-19, however no details of this group were available for analysis in this study [26].

The vaccine components itself include active immunising antigens, conjugating agents, preservatives, stabilizers, adjuvants, and culture media used in the preparation of the vaccine, which can be considered as potential triggers for an allergic reaction. Many delayed reactions are classified as Type III hypersensitivity reactions which are attributed primarily to the formation of immune complexes that including T cell-mediated processes. The most common signs of delayed-type reactions include rashes, which may include urticaria, erythema and angioedema [27,28].

The limitations of this study include the small number of potential thrombotic adverse events and the limited safety data currently available for the vaccine. There may be under-reporting by healthcare professionals and patients; the quality of reports may play a significant role that may cause false negative or false positive safety signals. Many confounders and incomplete data together with reporting biases may also limit the generalisability of the findings [13].

However, this study has provided valuable information about this current topical area from a trusted database. It has identified a very low number of investigated adverse reactions linked to the AstraZeneca vaccine that could be attributed to multiple causative factors and not merely the vaccines. The EMA’s Pharmacovigilance Risk Assessment Committee (PRAC) met on 18 March 2021. It concluded that the AstraZeneca vaccine was safe, effective and the benefits outweighed the risks and so urged people to accept the vaccination when offered [29].

Conducting further analyses based on more detailed thrombotic adverse event reports, including patients’ characteristics and comorbidities, may enable assessment of the causality with higher specificity. It must be undertaken by experienced personnel to identify any real risks associated with any and all COVID-19 vaccines.

## 5. Conclusions

This study has identified only 28 thrombotic events linked to the AstraZeneca vaccine out of 54,571 adverse reactions reported on the EV database. This finding should be interpreted cautiously as underreporting together with reporting biases may limit the generalisability of the findings.

It is difficult to determine a causal effect of the vaccine on the number of thromboembolic diseases reported. No clear causal effect can be confirmed, and multiple causative factors for thrombotic events were untested and undetermined. Further research is required to help national vaccine advisory boards and vaccine resistant people to make better informed, evidence-based decisions.

## Data Availability

The datasets generated during and/or analysed during the current study are available from the corresponding author on reasonable request.

